# MONITORING SARS-COV-2 TRANSMISSION AND PREVALENCE IN POPULATIONS UNDER REPEATED TESTING

**DOI:** 10.1101/2021.06.22.21259342

**Authors:** Matthew Wascher, Patrick M. Schnell, Wasiur R. Khudabukhsh, Mikkel Quam, Joseph H. Tien, Grzegorz A. Rempała

## Abstract

We describe a repeat SARS-CoV-2 testing model for monitoring and containing outbreaks in a residential community. The analysis is motivated by the Ohio State University (OSU)’s approach to monitoring disease at its Columbus, Ohio campus during the COVID-19 epidemic in autumn 2020. The model is simple, yet flexible enough to accommodate changes in behavior over time and to eliminate bias due to a nonrandom testing scheme. Model parameters are estimated from individual results of weekly SARS-CoV-2 testing of residents. Model output serves several purposes, including estimating the effective reproduction number and monitoring prevalence to help inform isolation and quarantine bed capacity. An extended version of the model is also considered where the residential population (on-campus students) is assumed to interact with another population for whom the testing regime is more relaxed and possibly less frequent (off-campus students or instructional faculty and staff). To illustrate the model application, we analyze both the synthetic data as well as the actual student SARS-CoV-2 testing data collected at OSU Columbus campus.

## 1. Introduction

In response to the COVID-19 pandemic, most American colleges and universities suspended on-campus residence and instruction in early March 2020, opting instead for predominantly online instruction (Rapanta et al., 2020). During the following summer, universities faced the decisions of whether and to what extent to resume on-campus residence and instruction, and, if they were to be resumed, what measures needed to be put in place to protect the health and safety of students, faculty, staff, and the surrounding communities (Smola, 2020). Several modeling and evaluation approaches were developed to assess the feasibility and impact of potential mitigation strategies such as frequent, random testing of asymptomatic individuals, contact tracing and isolation, caps on in-person class sizes. Some notable approaches involved full-scale agent- or network-based simulations parameterized using information from local epidemics and run forward to yield predictions through the end of an academic term, and thus naturally faced challenges stemming from a lack of transmission data in the campus setting and quickly increasing uncertainty in predictions of future trajectories, see, e.g., Christensen et al. (2020); Gressman and Peck (2020); Panovska-Griffiths et al. (2020); Chang, Crawford and Kaplan (2020). Nevertheless, these studies were able to robustly highlight the need for large-scale, frequent, randomized (if not comprehensive) testing of asymptomatic individuals (Gressman and Peck, 2020; Losina et al., 2020), which was adopted by many colleges and universities that ultimately held in-person instruction during the subsequent autumn semester.

The Ohio State University (OSU) partially resumed in-person instruction for the autumn 2020 semester, and implemented a battery of strategies to control COVID-19 incidence among associated individuals, especially students. The university conducted weekly routine PCR-based screening for SARS-CoV-2 among all (mostly undergraduate) students living on campus. In addition to aiding in isolation and contact tracing, trends in these data yielded insight into whether incidence on campus was increasing, decreasing or plateauing. The regular (but less frequent) random testing of asymptomatic undergraduate students living off campus in Columbus, OH was also initiated later in the semester with the primary intent of yielding information about incidence patterns off-campus that could then be used to inform mitigation strategies.

Motivated by the experiences of monitoring the spread of COVID-19 among undergraduate students at OSU on its main campus in Columbus, we present here a simple yet powerful modeling framework for the analysis of student testing data similar to that used at OSU in autumn 2020. Although for the purpose of analyzing OSU data we are only concerned with a single on-campus (residential) student population, our framework is quite general and thus allows for incorporation of any additional populations of interest, for instance, the off-campus students in the surrounding area or the university faculty and instructional staff. In both cases the goal of the analysis remains the same, namely to assess the ability of a repeated testing program to control on-campus (residential) outbreaks over the course of a fixed time horizon such as a college semester.

As we demonstrate in the paper, the naive estimates based on a regular testing scheme— with each member of the monitored population tested once per testing period (e.g., a week) on a day of their choice—suffer an upward bias when estimating the disease prevalence among the non-quarantined population, e.g., the daily proportion of tests with positive results. Since this bias increases in magnitude over the course of the testing period and also changes over the course of the epidemic, the data analysis of repeated testing needs to be conducted with extra care. This warrants a more complex, model-based approach that properly adjusts for the testing scheme effect. One possibility is to borrow the modeling framework from the classical epidemiology of infectious diseases and that is the strategy pursued in the current paper.

In order to describe our approach we first focus on a single (or closed) population model and then describe its extension to an open population case where we consider two or more interacting populations. The closed population model is used below to analyze OSU partial student SARS-CoV-2 testing data from the autumn of 2020. Despite sharing some similarities with recently proposed mechanistic models of SARS-CoV-2 repeated testing (see, e.g., Chang, Crawford and Kaplan 2020 or Paltiel, Zheng and Walensky 2020), it is, to our knowledge, the first detailed statistical model to analyze such data.

The underlying epidemiological framework for both closed and open models is a susceptible-exposed-infectious-removed (SEIR) compartmental process where changes over time are allowed in parameter values governing contact patterns, transmissibility, and social distancing. Therefore, the framework is appropriate for monitoring changes in underlying epidemic dynamics due to, for instances, the increase in virulence of the disease, the varying levels of compliance with social distancing measures or changing population vaccination levels. The basic SEIR framework is discrete, as it describes the evolution of the daily number of infections and is reminiscent of the classical Reed-Frost model of an epidemic (see, e.g., Andersson and Britton 2012). Both closed and open models are assumed to be informed by the results from some testing regimen implemented in their populations and fitted using the so-called dynamical survival analysis (DSA) approach based on the inferential methods described in Bastian and Rempala (2020); KhudaBukhsh et al. (2020); KhudaBukhsh et al. (2021), implemented here within the Bayesian framework via the Metropolis-Hastings-within-Gibbs (MH-within-Gibbs) sampler. This Bayesian approach to parameter estimation allows in particular for proper uncertainty quantification of the individual-level testing data. As these repeat testing data analyzed here may be considered interval-censored, we refer to the overall approach as the *interval* DSA or IDSA.

The model-based estimates are used to both track the effective reproduction number and provide projections supported by sequentially analyzed testing data – for use in decision-making about contact tracing and isolation capacity. Additionally, the estimates also help assess intervention efficacy and provide short-term forecasts of key quantities such as disease prevalence and quarantine/isolation (Q/I) resources needed. Although, as we show below, both closed and open population models avoid the upward bias associated with naive estimates of daily prevalence induced by the testing scheme, they have limitations, due to assumed complete or partial isolation of the residential population from the surrounding communities and due to ignoring social network structure and heterogeneity in activity patterns in the tested populations. In their current forms, the models also do not take into account the sensitivity and specificity of the PCR-based testing.

The remainder of the paper is organized as follows. In Section 2 we formulate both closed and open versions of the model and describe their statistical analysis based on the MCMC algorithm for the parameter estimation and validation. In particular, Sections 2.4 and 2.7 describe data sources and the process of integrating data into our models.

Section 3 presents a simulation study illustrating the model’s performance. We also briefly comment on the severity of bias in prevalence estimates based on naive counting methods such as Poisson regression – a popular tool in modern epidemiology and clinical research (Kianifard and Gallo, 1995) that could be considered an alternative to IDSA. In Section 3.5 we apply the closed model to data from weekly testing of OSU residential students in autumn 2020. Some concluding remarks are given in Section 4. The summary of notation as well as additional calculations on extensions of the open population model to *k* groups are provided in the appendix.

## 2. Methods

### 2.1. Data structure

We assume that repeated testing is performed in two coupled populations, for convenience referred below as off- and on-campus students. Key assumptions motivating our statistical model are that in the on-campus population:

1. All individuals are tested repeatedly (e.g., weekly), each time on a day of their choice within the fixed testing window.
2. The window between consecutive tests is shorter than the natural recovery time of the disease.

The testing schedule among the off-campus students is assumed to be less frequent and may not satisfy the above assumptions (see Section 2.6 below). In either population of students, we assume to know all individual testing dates and testing outcomes (positive or negative for SARS-CoV-2). A number of organizations (e.g. sports leagues, universities, etc) have used such testing schemes in response to the COVID-19 pandemic (Maloney, 2021; Walke, Honein and Redfield, 2020). OSU tested and collected data from August 8 to November 24, 2020 as part of the university’s plan to manage and mitigate COVID-19 cases during the fall semester with students on campus (OSU Monitoring Team, 2021). OSU’s policy was to test each of *n* = 12, 567 students living on campus weekly, keeping in mind that the natural recovery time for COVID-19 is thought to be between 10 and 14 days for the majority of cases. In the event of a positive test, the student was isolated for 10 days from the date of the test, in a designated Q/I location on campus, and contact tracing was initiated. Individuals identified as close contacts of a positive testing individual were quarantined for 14 days. We observed that in most cases there was a 2 day delay between administering the test and quarantining positive testing students. Students who tested positive and were subsequently quarantined were removed from the testing pool for the next 90 days.

The OSU COVID-19 surveillance dataset considered here consist of the dates of all tests for all students in the on-campus population, as well as the outcomes of those tests. Accordingly, with such data, we know the number of daily administered tests as well as daily testing positivity and negativity. The number of daily tests at OSU along with corresponding daily positive count is presented in Figure 1. As per institutional policy, we depict only data from 11950 students who were at least 18 years old at the time of testing. As we may see from the plot, at least for a portion of the time interval, the volume of daily testing on the first fours days of the week appears reasonably similar across different weeks. The corresponding volume of testing in the later part of the week is seen to be considerably lower.

**Fig 1:**
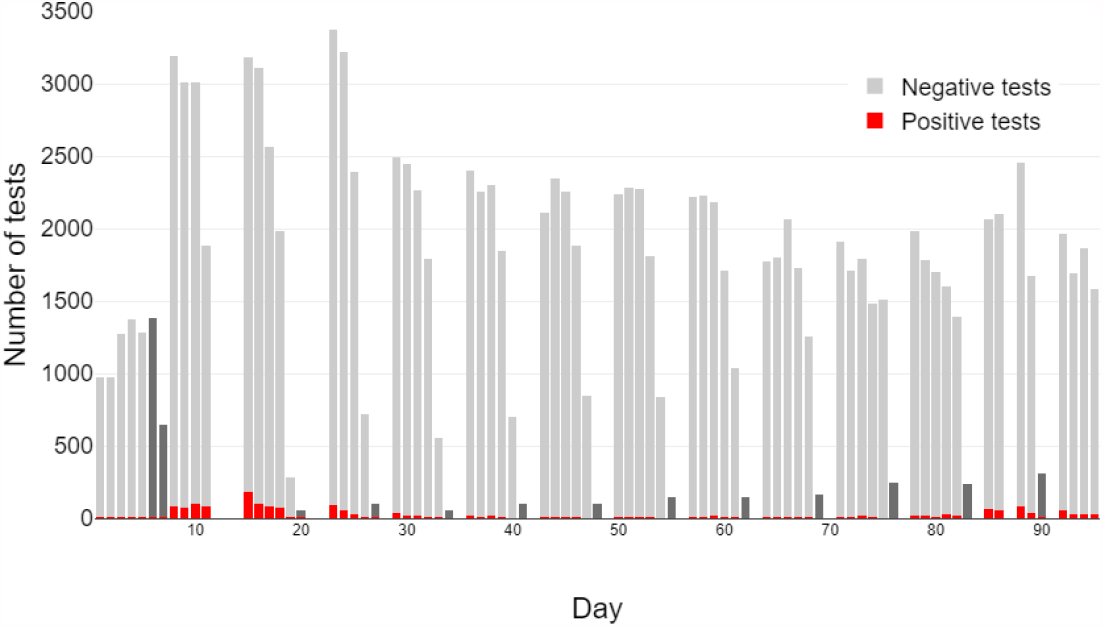
Testing volume for SARS-CoV-2 at OSU. The bars represent daily number of tests administered to residential OSU students (18 or older) in autumn 2020 colored by testing result. Weekend testing volumes were typically lower (often zero on Sundays) and are shown in darker gray color.

### 2.2. Closed model

As already indicated, the framework of our statistical inference model is based on the classical Reed-Frost type SEIR model of an epidemic spread (see, for instance, chapter 2 in Andersson and Britton 2012). This model separates individuals in a population into four categories of susceptible (*S*), infected or exposed (*E*), infectious (*I*), and removed (*R*). The exposed are assumed to be infected but not yet infectious and the removed cannot be reinfected. We treat our population as closed, well-mixed, and of known size. Below, we denote the initial susceptible population by *n*. Time is treated as a discrete, regular grid with units of days. We denote the counts of individuals in different categories (or compartments) at time *t* by *S*_*t*_, *E*_*t*_, *I*_*t*_, and *R*_*t*_ and assume that they evolve according to the following rules.

- Each pair consisting of one individual from *S*_*t*−1_ and one from *I*_*t*−1_ has probability *β*_*t*_(*n*) of infectious contact, and each individual among *S*_*t*−1_ who experiences such a contact becomes infected/exposed starting at *t*.
- Following infection/exposure, a susceptible individual enters the exposed (*E*) compartment. Each individual among *E*_*t*−1_ becomes infectious beginning at *t* (moves to *I*_*t*_) with probability *ψ* independently.
- Each infectious individual among *I*_*t*−1_ is removed beginning at *t* (enters the recovered (*R*) compartment) with probability *γ*_*t*_(*n*) independently.

We assume that individuals in *E* are not yet detectable by RT-PCR testing as SARS-CoV-2 positive. By contrast, individuals in *I* are both infectious and detectable. While the time from initial infection (exposure) to detectability (*ψ*^−1^) varies across individuals, it is typically shorter than five days (He et al., 2020; Larremore et al., 2021). Accordingly, we assume *ψ* = 0.25 in our simulation study and data examples below.

Let *δ*_*t*_ denote the daily decrease in the count of susceptibles and *ε*_*t*_ denote the daily decrease in the count of infectious individuals. From the above discussion we derive the following probability laws for the daily increments of infection *δ*_*t*+1_ = − (*S*_*t*+1_ − *S*_*t*_) and recovery *ε*_*t*+1_ = *R*_*t*+1_ − *R*_*t*_, respectively:

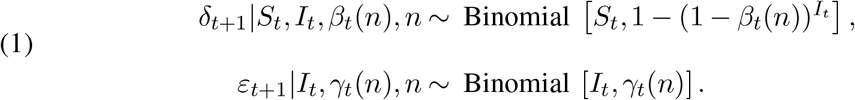

The model parameters are therefore the rates of infection transmission *β*_*t*_(*n*), and of removal *γ*_*t*_(*n*), as well as the initial conditions *S*_0_, *E*_0_, and *I*_0_ (*R*_0_ = 0). We assume in the analysis that the initial conditions are known, and estimate *β*_*t*_(*n*), *γ*_*t*_(*n*) from data. Note that recovery here encompasses not only biological clearance of infection, but also removal from infectiousness due to isolation following positive test, as well as removal due to quarantine of infected contacts of positive cases.

An important feature of the model is that both *β*_*t*_(*n*) and *γ*_*t*_(*n*) parameters depend on *t* and can potentially change at multiple time points. The model thus can accommodate behavioral changes over time, for example in response to perceived risk of infection, policy changes, weekend or holiday effects, and more.

### 2.3. Survival and hazard functions

As indicated in the introduction, the estimation approach builds on the DSA method given in KhudaBukhsh et al. (2020). Specifically, we adapt the method to individual-level repeat testing data. There are several advantages of applying DSA to our setting such as the automatic correction for the interval censoring and the testing bias, both introduced by the data structure assumptions given in Section 2.1. See discussion in Section 3.1 for further details.

Consider the survival function 𝒮_*t*_ that describes the decay of susceptibles over time, along with its associated hazard function *h*_*t*_. More precisely, 𝒮_*t*_ is the probability that an initially susceptible individual is still susceptible at time *t*. Define *β*_*t*_(*n*) = *β*_*t*_*/n* when *n* is assumed to be large (i.e., we have a large initial population of susceptibles) and further assume that *γ*_*t*_(*n*) = *γ*_*t*_ is *n*-free. The probability that an initially susceptible individual stays susceptible until *t* is given by

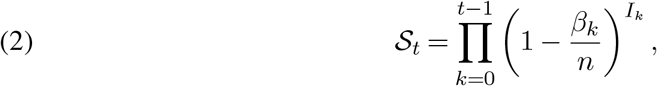

and thus the hazard function for a random susceptible being infected in [*t, t* + 1] is

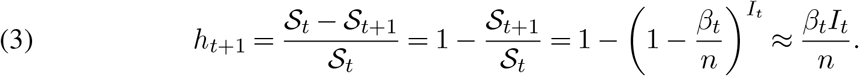

Since by a similar calculation the hazard of recovery in the interval [*t, t* + 1] is seen as simply *γ*_*t*_, we may consider the simple approximation to (1):

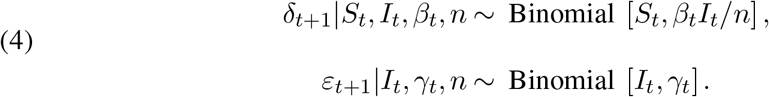

We also note that given the values *S*_*t*_, *β*_*t*_, *γ*_*t*_, and *n*, we have the following expression for the *effective reproduction number* at time *t*:

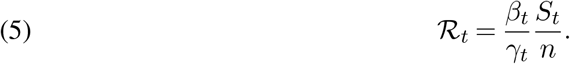

As the *E* compartment is not observed (since individuals in *E* do not test positive – see Section 3.3), it is useful to consider the decay of the combined count of susceptible and exposed individuals. To this end, consider an initially susceptible individual. The probability that this individual has not yet entered *I* by time *t* can be computed by convolving over the transitions from *S* to *E* and *E* to *I*, giving

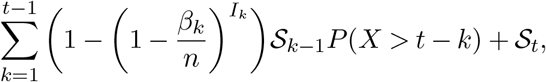

where *X* ∼*Geom*[*ψ*]. The probability that an initially exposed individual has not yet entered *I* by time *t* is given be *P* (*X* > *t*) = (1 − *ψ*)^*t*^. Now consider the probability that a randomly chosen individual from among those initially either susceptible or exposed has not yet entered *I* by time *t*. Using the above along with (3) we may approximate this probability by

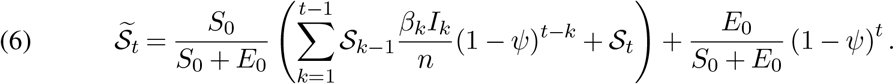

### 2.4. Testing results and infection/exposure times

Since each on-campus student undergoes weekly SARS-CoV-2 testing (we assume here that the noncompliance effect is negligible, this was the case at OSU) for each individual we have available

- *t*_*neg*_, the most recent time this individual was known to be susceptible or exposed, and
- *t*_*pos*_, the first time this individual was known to be infectious.

Note that it is possible that a particular individual was infectious the first time they were observed, in which case we set *t*_*neg*_ = 0. It is also possible that a particular individual has never been observed to be infectious, in which case we set *t*_*pos*_ = ∞.

Given 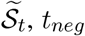, and *t*_*pos*_, we can find the probability that an individual became infectious on a particular day as follows:

- If *t*_*neg*_ = *i* and *t*_*pos*_ = *j*, then for each *i* < *k ≤j*, the probability that this individual became infectious on day *k* is

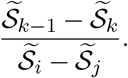
- If *t*_*neg*_ = 0 and *t*_*pos*_ = *j*, then for each *i* < *k ≤j*, the probability that this individual became infectious on day *k* is

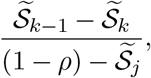

where *ρ* = *I*_0_*/n*.
- If *t*_*neg*_ = *i* and *t*_*pos*_ = *∞*and we have observed data until present time *T*, then for each *i* < *T* the probability that this individual became infectious before time *T* is

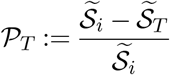

and thus the probability this individual became infectious on day *k* with *i* < *k ≤ T* is

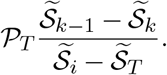

### 2.5. Closed model estimation

We devised an iterative procedure implemented via the MH-within-Gibbs sampler algorithm to estimate the model parameters. Following initialization, we use the current prevalence estimate to compute the survival function (2). The survival function and individual interval censored testing data are then used to compute daily incidence, which is then used to update the prevalence estimate. We assume that each exposed individual *i* remains exposed for *X*_*i*_ days where *X*_*i*_ ∼Geom [*ψ*] before moving to the *I* compartment. Below we denote the number of arrivals into *I* on day *t* by *ω*_*t*_ and the realizations of the random variable *X*_*i*_ by *x*_*i*_ for *i* = 1, …, *n* where *n* is the number of initially susceptible. The detailed Gibbs sampler algorithm is as follows.

1. Initialize 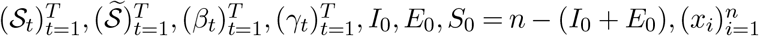.
2. Initialize 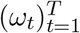 by drawing each individual infectious time (entering of the *I* compartment) from

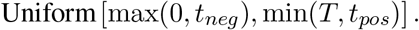
3. Given the number of infections, calculate 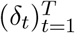 by drawing each individual infection time (entering of the *E* compartment). Given previous set of draws 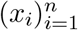, propose each new 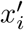 as an iid draw from Geom [*ψ*] and accept with probability

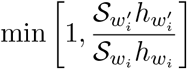

where 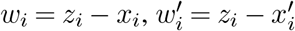 and *z*_*i*_ denotes the recovery time of the *i*-th individual. See Section B.1 in the appendix for the derivation of this probability.
4. Given 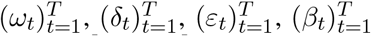, and 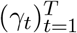,propose 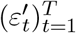 by drawing each 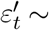 ∼ Binomial 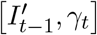 independently, where 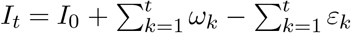 and 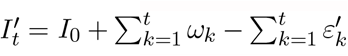. Accept the vector 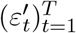 with probability

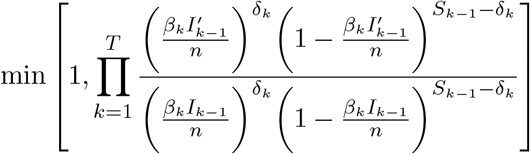

(see Section B.2 for the justification).
5. Given 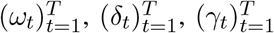,and 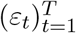 calculate 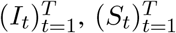,and 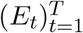 by using

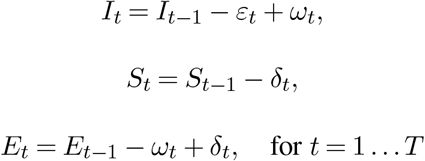
6. Update 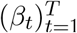 by drawing β_*t*_ *I*_*t*_/*n* ∼Beta[*δ*_*t*_ +*a, S*_*t*−1_ − *δ*_*t*_+*b*].
7. Update 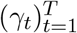 by drawing γ_*t*_ ∼Beta[*ε*_*t*_ +*c, I*_*t*−1_ − *ε*_*t*_ +*d*].
8. Given 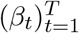 and 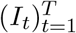, update 𝒮_*t*_ using (2).
9. Given 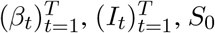 and *E*_0_, update 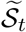 using (6).
10. For each individual, compute the probability of infectious time occurring on a specific day as described in Section 2.4 and update 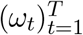 by drawing the infectious time from this distribution for each individual.
11. Go to step 3 and repeat until convergence.

The above estimation scheme yields posterior samples for 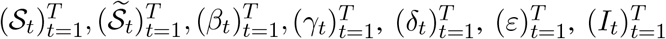, and 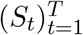. Note that the updating 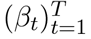 and 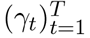 here uses a Beta-Binomial conjugate prior model. For the transmission parameters *β*_*t*_ we have the prior Beta [*a,b*], and for recovery parameter *γ*_*t*_ we have the prior of Beta [*c,d*]. In our simulation study and data example, we choose *a* = *b* = 1 and *c* = 10, *d* = 50, resulting in an uninformative prior for *β*_*t*_ and a strongly informative prior for *γ*_*t*_. The choice of a strongly informative prior for *γ*_*t*_ reflects the fact that the testing scheme governs most removals and therefore gives us much information about *γ*_*t*_. In fact, in the idealized version of the scenario in 2.1, we know all the removal times exactly and could use them directly. However, since the model is unlikely to hold perfectly in practice, an indirect, parametric approach with a strong prior is preferred, as it simultaneously allows us to utilize the empirical information about removals and to account for uncertainty due to small violations of the assumptions described in Section 2.1. The choices of *c* = 10 and *d* = 50 reflect the fact that under a weekly testing scheme, the expected time to removal is 6 days.

We note that the Metropolis steps in steps 3 and 4 both have acceptance ratios close to 1, suggesting the proposal distributions are close to the full conditionals. This means the MCMC mixes reasonably quickly. We also note that step 10 is in fact an approximation as the collection 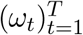 is not independent in the full conditional. However, in KhudaBukhsh et al. (2020) it was shown that this collection is independent in the limit as *n →∞* and we use this fact to justify the approximation. One could also consider using a Metropolis-Hastings step here, but we found that in practice it is difficult to generate good proposals ensuring fast mixing and that the limiting approximation is generally satisfactory.

### 2.6. Open model

The closed model in Section 2.2 considers a single population in isolation, with residential undergraduates as a motivating example. However, in practice this may not be an isolated population, and interactions with other groups (e.g., faculty, staff, off-campus students, and local residents) may affect disease dynamics in the original population under study. These additional populations may not be undergoing repeated testing at the same frequency (or not at all), and as a result, less information is available about them. To address this issue, we consider an extension of the initial model to include an “off-campus” population as discussed in Section 2.1. We assume that although we are no longer in control of the testing process in this second population, its infection prevalence and population size estimates are nevertheless independently available through some different sampling efforts (for instance, by a state agency or employee healthcare provider). For simplicity, in our simulation study below we also assume that, in the time window of interest, the total off-campus population is known and the external infection rates is constant.

Our proposed “open” model in this setup is as follows. Each of the 2 groups of interest (on- and off-campus populations) is divided into SEIR compartments as before. We denote the total numbers of individuals in each group by *n*_1_ and *n*_2_ respectively, the numbers of susceptible individuals at time *t* in each group by 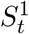 and 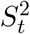 respectively, the numbers of infected individuals in each group at time *t* by 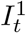 and 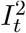 respectively, and so on. The compartments then evolve according to the following rules:

- Each pair of one individual among 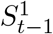 and one among 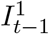 has probability 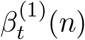 of infectious contact, and each individual among 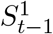 who experiences such a contact is infected/exposed beginning at *t*.
- Each pair of one individual among 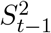 and one among 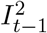 has probability *β*^(2)^(*n*) of infectious contact, and each individual among 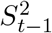 who experiences such a contact is infected/exposed beginning at *t*.
- Each pair of one individual among 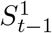 and one among 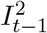 has probability *β*^*∗*^*/n*_1_ of infectious contact, and each individual among 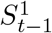 who experiences such a contact is infected/exposed beginning at *t*.
- Each pair of one individual among 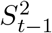 and one among 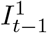 has probability *β*^*∗*^*/n*_2_ of infectious contact, and each individual among 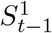 who experiences such a contact is infected/exposed beginning at *t*.
- Following infection/exposure, an individual enters the exposed compartment. Each individual among 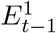 and each individual among 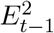 moves to the corresponding *I* compartment at time *t* with probability *ψ* and then becomes infectious.
- Each infectious individual among 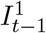 is removed beginning at *t* with probability 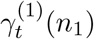.
- Each infectious individual among 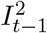 is removed beginning at *t* with probability 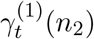.

Let 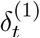 and 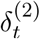 be the daily decrease in susceptible counts for groups 1 and 2 respectively, and let 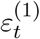 and 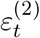 be the daily decrease in infectious counts for groups 1 and 2 respectively. The approximate probability laws for these quantities are as follows,^1^

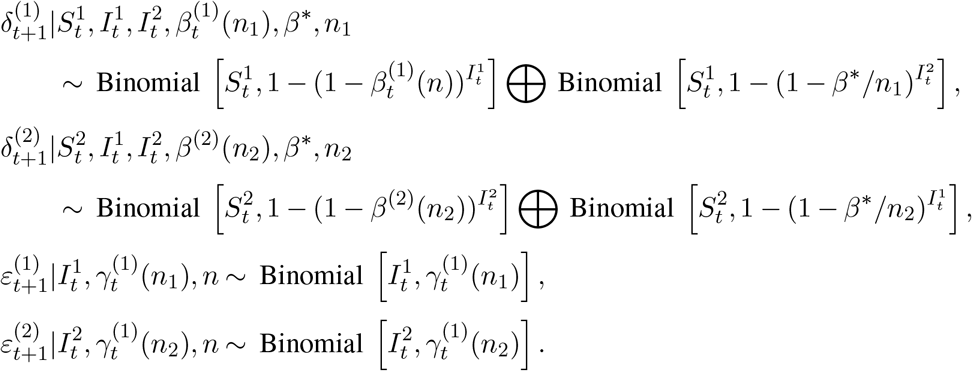

Define, as before, 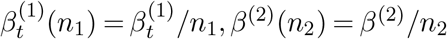 and 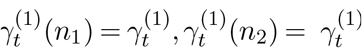. Note that assuming that *n*_1_ and *n*_2_ are sufficiently large, we may approximate the laws of 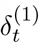 and 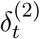 using Le Cam’s theorem (Le Cam et al., 1960).

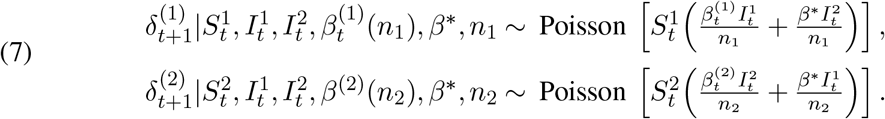

If we now let

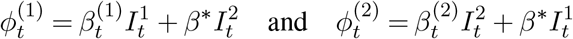

then we can rewrite (7) as

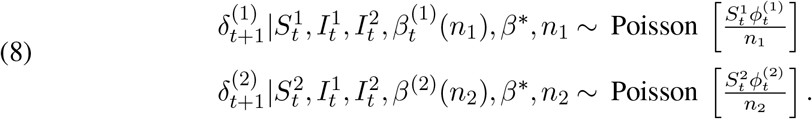

### 2.7. Two group survival functions

Let 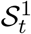 and 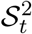 be the survival functions that describe the decay of susceptibles for groups 1 and 2 respectively. Conditional on 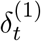 and 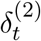, these can be described by the recursive equations

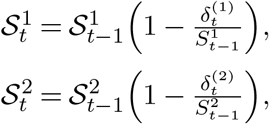

and so the marginal distributions are given by

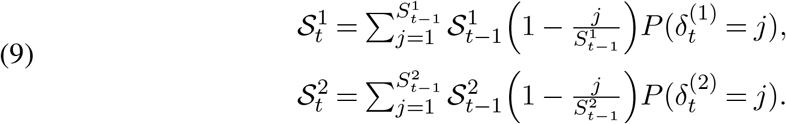

As before, to incorporate the *E* compartment (so only individuals in the *I* compartment can test positive), we define 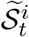 to be the probability that an individual randomly chosen from among those initially either susceptible or exposed has not yet become infected by time *t*. As before, let *X* ∼ Geom[*ψ*] so that for each *i* = 1, 2 this probability is given by

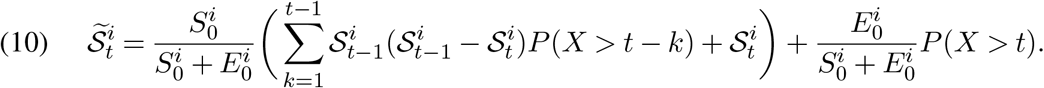

We also define the *partial* effective reproduction number denoted 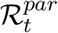 as follows

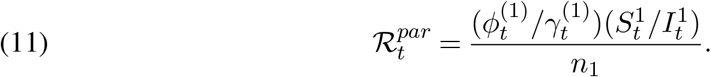

Note that this reproduction number is partial in the sense that it only pertains to the reproduction rate in the first group (on-campus population).

### 2.8. Open model estimation

In this setting, the data about the primary group is collected as in Section 2.1. Rather than assuming any specific testing regimen for the off-campus group, we assume instead that the available estimates of prevalence and population size *n*_2_ allow us to estimate 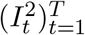. In addition, we also treat the values of *ψ* and 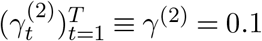 as known. Indeed, as *ψ* = 0.25 and *γ*^(2)^ correspond to the incubation and recovery period of the disease (in both on- and off-campus population), they are assumed to depend on inherent (and known) characteristics of the virus rather than any statistical properties of the sampled population or the sampling scheme itself. This removes the need to estimate 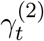 and 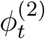. For 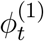 we use the Gamma-Poisson conjugate prior with prior hyperparameters *a*_1_, *b*_1_, and for 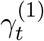 we use a Beta-Binomial conjugate prior with hyperparameters *c*_1_, *d*_1_. The choice of hyperparameters may depend on the disease and testing scheme; we discuss this below after first describing the algorithm as follows.

1. Initialize 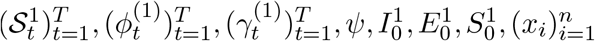.
2. Define 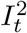 based on its estimated values.
3. Initialize 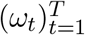 by drawing each individual infectious time (entering the *I*^1^ compartment) in group 1 from

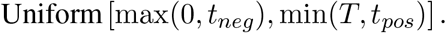
4. Given the number of infections, update 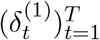 counts by first drawing each *i*-th individual infection time (entering of the *E* compartment) and then subtracting *x*_*i*_. Given each previous draw *x*_*i*_ propose new set of 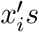 as an iid draw from Geom [*ψ*] and accept it with probability

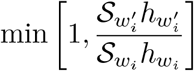

where 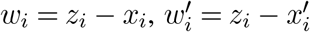 and *z*_*i*_ is the recovery time for the *i*-th individual. See Section B.1 in the appendix for the derivation of this probability.
5. Given the other parameters, draw 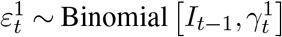 and calculate

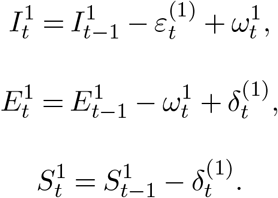

for *t* = 1, …, *T*.
6. Given the other parameters, draw for *t* = 1, …, *T*

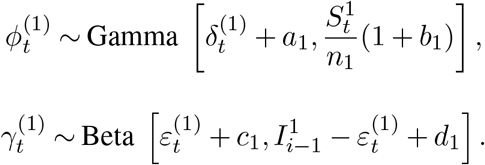
7. Given the other parameters, update 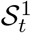 recursively according to (9).
8. Given 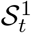, update 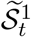 according to (10).
9. For each individual, compute the probability of infection time occurring on each day as described in Section 2.4 and update 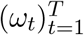 by drawing the infection time from this distribution for each individual.
10. Go to step 4 and repeat until convergence.

When using the above algorithm in our synthetic data analysis, we have made several adjustments to enhance its performance. Because much of the short term fluctuation in 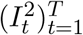 is likely due to randomness in the testing sample, we replaced 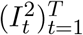 with a spline-smoothed version of itself. For *ϕ*_1_ we used a diffuse prior setting *a*_1_ = *b*_1_ = 0.01. Finally, we let *c*_1_ = 10, *d*_1_ = 50 and thus used a strong informative prior centered at 1*/*6 for 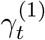. As in the closed model, this choice seems justified by the information contained in the testing scheme and is reflective of the 6 day average time to removal.

The extension of the current open model to include additional on-campus populations (groups) is relatively straightforward and although we do not pursue it in this paper, the outline of necessary derivations is provided in the appendix.

#### 2.8.1. Estimating *β*^∗^

One of the advantages of the open model described in Section 2.6 is that with some additional effort we can also estimate the cross-infection rate *β*^*∗*^. Although in the framework given in Section 2.8 *β*^*∗*^ is not identifiable if the sequence 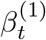 is assumed to change every day, that assumption may be often relaxed.

For instance, assuming a ‘shelter in place’ order was issued for a time interval of at least two days [*t*_1_, *t*_2_] *⊂* [0, *T*], it is reasonable to expect that 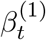 is constant 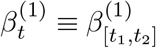 on this interval. In this case we may obtain posterior samples of *β*^*∗*^ via an additional step performed after step 10 in the previous section Gibbs sampler, where we use the current values of *ϕ*^(1)^, *I*^1^, and *I*^2^ over the interval [*t*_1_, *t*_2_] to perform the least squares projection on the overdetermined system

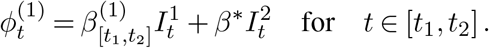

yielding posterior samples of *β*^*∗*^. As an alternative, we could also use the above relation to directly obtain the estimating equations for *β*^*∗*^ and 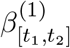 based on the posterior estimates of *ϕ*^(1)^, *I*^1^, and *I*^2^ averaged over [*t*_1_, *t*_2_].

## 3. Results

### 3.1. Bias in naive, non-mechanistic modeling

Before we present the proposed IDSA model performance on data examples, let us first briefly discuss the alternative non-mechanistic approaches often considered in similar contexts in the statistical literature. Indeed, by now a plethora of approaches to analyzing COVID-19 occurrences via count-regression models both in the open and closed population settings have been proposed (see, e.g., Chan et al. (2021); Kianifard and Gallo (1995) and references therein). In our current setting of repeated testing, it is perhaps especially tempting to entertain a simpler and more intuitive alternative to IDSA, by modeling the epidemic using daily counts of tests administered and daily positivity. However, the sample of individuals tested each day is not random and depends on the specific testing regime. This could potentially cause bias and adversely affect any planned intervention. To give a simple example, we simulated 1000 epidemic trajectories using the same settings as in Section 3.3 below, comparing the naive estimate of prevalence computed using the formula (daily positivity rate * (*n* −*R*_*t*_)) with the true value of *I*_*t*_, and averaged over the 1000 simulations. The results are presented in the left panel of Figure 2, illustrating a positive bias in the implied prevalence that increases with the true *I*_*t*_ and also over the course of the week (the duration of testing window). Indeed, note that for any day in the week-long testing window beyond day one (*d* > 1), the daily positivity rate (i.e., the daily proportion of positive tests) has to overestimate the prevalence, since individuals in the tested sample will test positive so long as their infection time is smaller than *d*. Since an individual’s test day is chosen uniformly at random from the days of the week, it is approximately independent of that individual’s infection day so that the bias gets accumulated over the testing window. In Figure 3 we demonstrate this mechanism for day two of testing (*d* = 2).

**Fig 2:**
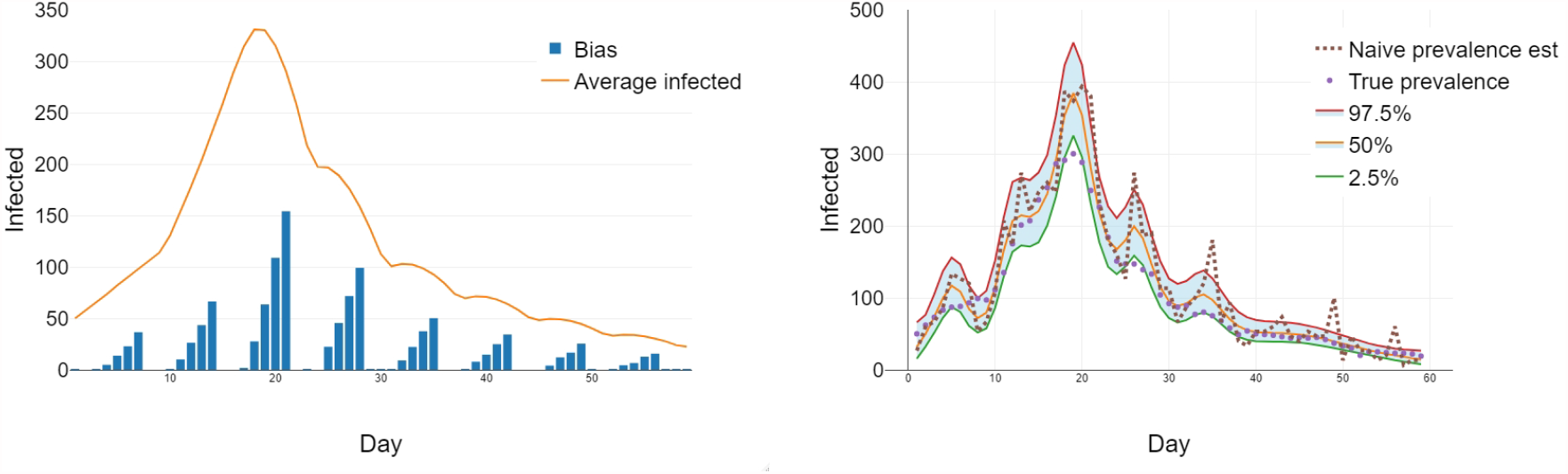
Bias in testing-based estimates of prevalence. *Left:* The blue bars depict average differences between daily estimates of prevalence computed from daily positivity rate and the true values of *I*_*t*_ (yellow line) averaged over the 1000 simulations. The bias is seen to increase in the course of the week and with true *I*_*t*_. The mechanism of the bias is explained in Figure 3. *Right*: Poisson GAM fit (solid lines) of the prevalence along with the naive prevalence estimate (brown dotted line) obtained by *n ∗* (daily positivity) compared to the actual simulated counts (purple dots). The Poisson model is seen to severely overestimate *I*_*t*_ following the pattern of bias from the left panel.

**Fig 3:**
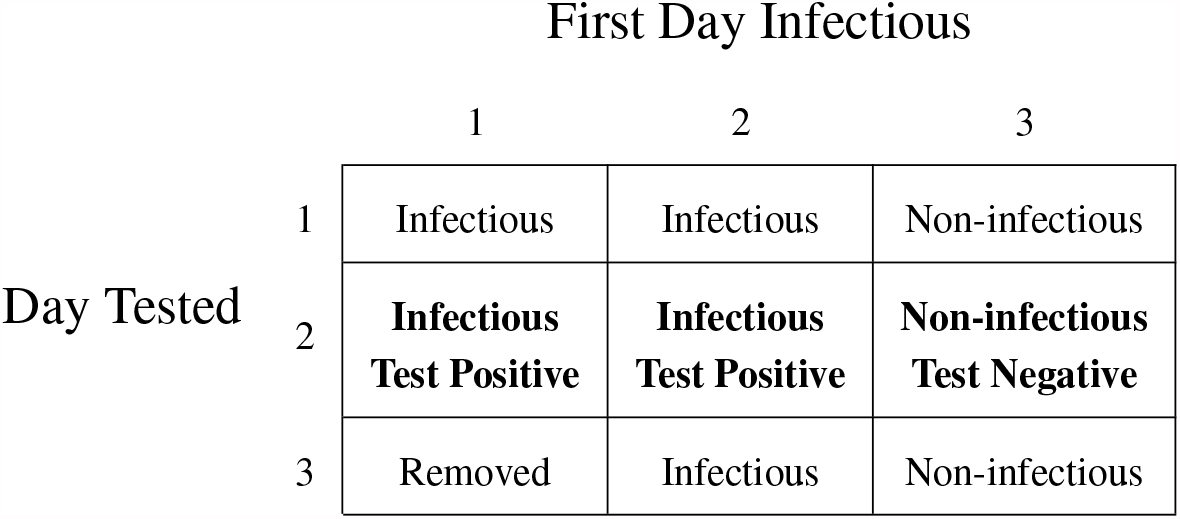
Example of bias mechanism for test day d = 2. Assuming that test day is independent of infection day, we define prevalence (Pre) as the proportion of true infectious in non-removed population. If we define the positivity (Pos) as the proportion of infected in the sample tested on day *d* = 2 then it is readily seen that Pre < Pos, creating a bias. This bias may be quite substantial, depending on the epidemic dynamics, as seen in Figure 2.

In order to examine the implications of the positivity bias on a modeling strategy relying on daily incidence counts, the SEIR model and testing data were simulated (see Section 3.3 for details) giving us the hypothetical daily number of tests administered along with the daily positivity rate. Under the assumption that removals of infectious individuals are deterministic (occurring only at a fixed time after a positive test, and with no untested recoveries), we calculated the daily number of individuals in the S, E, or I compartments and fitted the Poisson generalized additive model to the simulated dataset. The model used the number of positive tests as the outcome variable, the log number of tests administered as the offset, and a penalized spline term for time via an adaptive spline basis (bs = “ad”) with otherwise default settings of the version 1.8-33 of R `mgcv` package function `gam()`. Prevalence estimates were then produced at each timepoint (day) by predicting the number of positive tests, if the entire non-removed population had been tested (by setting the offset to the log number of non-removed students). The results are shown in the right panel of Figure 2. As can clearly be seen from the plot, the Poisson model substantially overestimates the peak of the epidemic and exhibits large oscillations along the epidemic trajectory that are consistent with the accumulated bias in the naive prevalence estimates. We thus see the naive Poisson model is in fact attempting to fit (with some smoothing) these incorrect, overestimated prevalence values.

### 3.2. Simulation study: Overview

In order to assess the performance of the estimation algorithms proposed in Sections 2.5 and 2.8, we performed a simulation study for both the closed and the open models. The parameter values for the simulations were chosen in a way that produced epidemic curves that were qualitatively similar to what was observed over the first 60 days at the Ohio State University Columbus campus: an early peak followed by a decline and a plateau. R code used for these simulations can be found as https://github.com/mwwascher/IDSA.

### 3.3. Simulation study: Closed model

For the closed model, we considered an SEIR epidemic in a population of size *n* = 10000 over a period of time *T* = 60 days. In order to account for changing social behavior and compliance with public health measures, we let *β*_*t*_ vary over the course of the epidemic, setting *β*_*t*_ = .35 for *t* = 1, …, 7, *β*_*t*_ = .45 for *t* = 8, …, 14, *β*_*t*_ = 0.08 for *t* = 15, …, 28, and *β*_*t*_ = 0.12 for *t* = 29, …, 60. We assume that when a susceptible individual becomes infected they enter the *E* compartment, where on each day they become infectious and move to the *I* compartment with probability *ψ* = .25 independently. As already indicated earlier, we assume this probability is an inherent biological characteristic of the disease and is thus known. We set the initial numbers of susceptible, exposed, infectious, and removed individuals to *S*_0_ = 9900, *E*_0_ = 50, *I*_0_ = 50, *R*_0_ = 0.

We assume that each individual in the population is tested weekly. We simulate this each week by numbering individuals 1, …, 10000, permuting this sequence into a random order of individual tests distributed approximately uniformly over 7 days, Monday through Sunday. Individuals who test positive are removed 2 days after being tested, due to the delay in test outcomes reporting. Additionally, each positive test results in removing *X* ∼Geom[*p*] (where *p* chosen so that *EX* = 0.15) additional positive individuals from the *I* compartment through contact tracing. Thus, the only way to be removed from the *I* compartment is due to a positive test or contact tracing, since by assumptions in Section 2.1 these removals happen faster than the natural recovery from the disease.

For each individual, we record the time of that individual’s first positive test or time of contact tracing, if one exists. We additionally record either the time of the most recent negative test before the positive one, or if the individual never tested positive or was contact traced, simply the time of the the most recent negative test. This individual-level information constitutes data that is used to fit the model. For comparison, we also record the true numbers of *S, E, I*, and *R* individuals each day. These numbers are not used to fit the model but rather to assess the quality of model fit. We fit the model using the algorithm in Section 2.5 with a noniformative prior Beta[1, 1] for *β*_*t*_ and an informative prior Beta[10, 50] for *γ*_*t*_ centered at 1*/*6. The choice of this particular value is based on the mean time (in days) to removal under the weekly testing regime. We ran the MCMC for 10000 iterations, discarding the first 5000 as burn-in and retaining the second 5000 as posterior samples.

The results of the IDSA fit for the closed model are presented in the two panels of Figure 4. In the left panel, the median, the 2.5% and the 97.5% quantiles of the posterior estimates of the daily prevalence are plotted over the time window *T* = 60 days, along with the actual prevalence *I*_*t*_. The comparison of *I*_*t*_ with median model prediction indicates a reasonably good model fit, except perhaps for the first few days. Importantly, the IDSA model correctly identifies both the timing and the size of the epidemic peak and avoids the naive model positivity bias. We note that the IDSA plot may be directly compared with the Poisson regression plot in the right panel of Figure 2 since both models were fitted using the same simulated epidemic trajectory (the purple dots indicate the same true prevalence in both figures). The right plot of Figure 4 compares the empirical effective reproduction number ℛ_*t*_ (given in (5)) for the simulated trajectory with the IDSA model estimate summarized by the three quantile plots (median and 2.5% and 97.5% quantiles) of the posterior estimates. Again, it appears that there is a good agreement with the observed values of ℛ_*t*_, even at the locations (marked by vertical lines) where the true *β*_*t*_ values change, except for the initial time period. Indeed, for the initial few days the small amount of data and the delay in obtaining initial testing is seen to contribute to model bias in estimating ℛ_*t*_. Despite this initial bias though, we see from the left panel in Figure 4 that the IDSA closed model fits the synthetic data well, as it places the peak of the epidemic at the true location, and its posterior 95% credible interval covers the true prevalence values.

**Fig 4:**
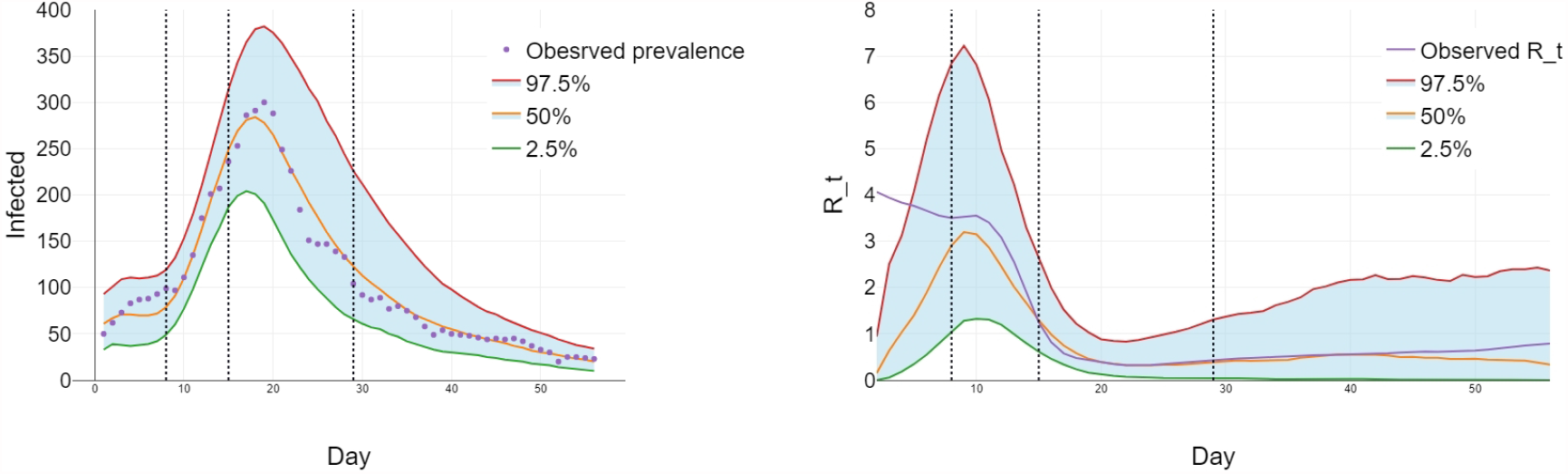
IDSA closed model fit. *Left:* The model estimates of prevalence represented by the posterior quantile plots (smooth curves) compared with the true observed prevalence of the simulated trajectory. The quality of IDSA fit may be directly compared to the fit of the Poisson model shown in the left panel of Figure 2. *Right:* Model-based posterior estimates of ℛ_*t*_ given by (5) compared to the true observed values calculated for the simulated trajectory.Vertical lines indicate the change points for the infection parameter *β*_*t*_.

### 3.4. Simulation study: Open model

For the open model consisting of two populations (on- and off-campus), we again consider an SEIR epidemic over a *T* = 60 day period. We begin by defining the first group (on-campus population) and its internal parameters in the same way as in Section 3.3, except that we set 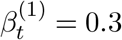 for *t* = 1, …, 7, 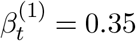 for *t* = 8, …, 14, 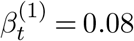 for *t* = 15, …, 28, and 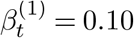 for *t* = 29, …, 60. We also add a second group (off-campus population) of size *n*_2_ = 30000. We set for this group, 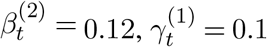 for all *t* = 1, …, 60. Additionally, we take 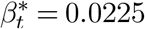 for all *t* = 1, …, 60 to be the rate of cross-infection of the first group by the second. As before, we assume that all individuals move from the compartment *S* to *E* upon infection (exposure) and may move from *E* to *I* with known probability 0.25 each day independently. Finally, we set the initial numbers of susceptible, exposed, infectious, and removed individuals to *S*_0_ = 29, 400, *E*_0_ = 300, *I*_0_ = 300, *R*_0_ = 0.

The testing schedule for the first group remains the same as in the closed model in the previous section. For group two, we assume that each individual is tested once every 30 days, and we generate the randomized testing schedule analogously to group one: each testing period we schedule tests in random order and uniformly over 30 days. No action is taken when an individual in group two tests positive and no contact tracing is performed in this group. Thus, the individuals (off-campus population) are assumed to leave the *I* compartment via natural recovery only. The prevalence estimate for group two is obtained as the scaled proportion of daily positive tests.

For group one, the model is given data as in Section 3.3. For group two, the model is given the number of positive tests out of the 1000 total daily tests for each day in the 60 day period. We then use the algorithm in Section 2.8 to fit the model. We used a diffuse Gamma[0.01, 0.01] prior for 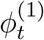 and an informative Beta[10, 50] prior for 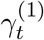. We ran the MCMC for 10000 iterations, discarding the first 5000 as burn-in and retaining the second 5000 as posterior samples. Recall that despite the model structure expansion, the first group is still of primary interest; indeed, the second group is only introduced as a “nuisance component” to account for the effects of the surrounding area and its populace on the first group. The results of the IDSA fit to the open model are presented in the two panels of Figure 5.

**Fig 5:**
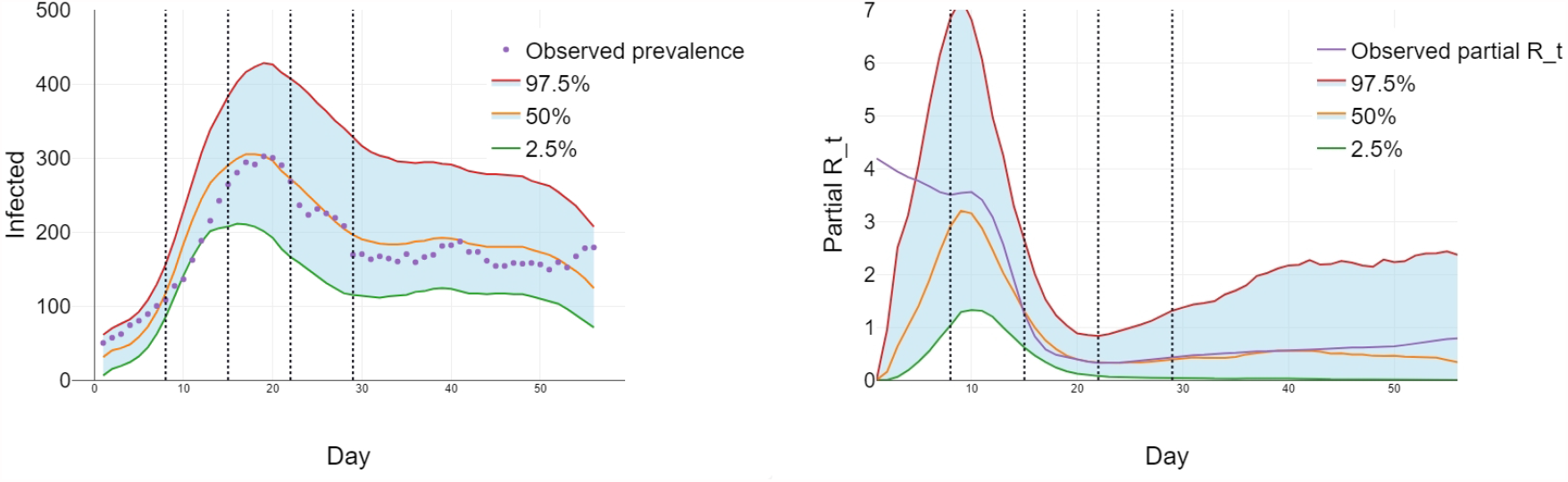
IDSA open model fit. *Left:* The model estimates of prevalence in the first group (on- campus population) represented by the posterior quantile plots (smooth curves) compared with the true observed prevalence of the simulated trajectory (purple dots). *Right:* Model- based posterior estimates of 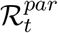 given by (11) compared to the true observed values for the simulated trajectory. Vertical lines indicate the change points for the infection parameter *β*_*t*_. The model estimated values of 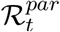 trail off at the end of the time window because some infections that would be imputed to this time period will have occurred after individuals’ most recent tests.

In the left panel, the median, the 2.5% and the 97.5% quantiles of the posterior estimates of the daily prevalence in the first group (on-campus population) are plotted over the time window *T* = 60 days, along with the corresponding actual prevalence in the first group 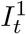. As we can see, despite the more complex simulated data structure than that analyzed in the previous section, the IDSA median model again makes a reasonable prediction and provides a good fit. As before, the IDSA model correctly identifies both the timing and the size of the epidemic peak and manages to avoid the testing positivity bias. The right plot of Figure 5 compares the partial effective reproduction number 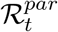 (given in (11)) for the simulated trajectory with the IDSA model estimate summarized by the three quantile plots (median, the 2.5% and the 97.5% quantiles) of the posterior estimates. Again, it appears that there is a good agreement with the true values of ℛ_*t*_ across the relevant time window including at the locations (marked by vertical lines) where the true *β*_*t*_ values changes. As in the previous section example, the IDSA fit places the peak of the epidemic close to the true peak, and the posterior 95% credible interval covers 96.4% of the true incidence values and 96.4% of the 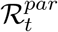 values, although the 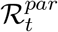 estimates trail off slightly at the end of the time period due to backfill (delayed results reporting, see caption in Figure 5). As with the close model, we observe also some bias in the initial the estimate of 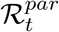 due to insufficient data.

We also estimated *β*^*∗*^ using the method described in Section 2.8.1, assuming that 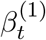 is constant on the subinterval from days 22 to 28 and that 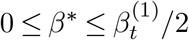 for all *t*. We obtained a posterior median of 0.022 and a 95% posterior interval of (0, 0.034) whereas the true value of *β*^*∗*^ was 0.0225.

### 3.5. Analysis of OSU COVID-19 surveillance data

In this section we present the data analysis that has motivated the IDSA model. We use the slightly altered^2^ surveillance dataset on student testing results collected by The Ohio State University between August 17 and November 24, 2020 as part of the university’s plan to manage and mitigate COVID-19 during the autumn semester 2020 with students on campus. The OSU’s internal policy was to weekly test each of *n* = 12, 567 students living on campus. In the event of a positive test, the student was isolated for 10 days and contact tracing was initiated. We observed that in most cases there was a 2 day delay between administering the test and quarantining positive testing students. Students who tested positive were subsequently quarantined and removed from the testing pool for 90 days. To analyze these data we consider a closed model only, since during a large part of the autumn semester 2020 only limited data were available about students living off-campus, an important component of the background group in the open model in this setting.

The OSU dataset consists of the date of the first known positive test result (if one exists) for each student living on campus and the date of the most recent negative test result before the positive test (or the most recent negative test result if no positive test result exists). In addition, the dataset contains information on the number of tests administered each day, and the daily number of positive and negative test results.

The IDSA closed model is fit to the data as in Section 3.3, by applying the MH-within-Gibbs sampler algorithm described in Section 2.5. We used a noninformative Beta[1, 1] prior for *β*_*t*_ and an informative Beta[10, 50] prior for *γ*_*t*_ centered at 1*/*6. We ran the MCMC for 10000 iterations, discarding the first 5000 as burn-in and using the remaining to approximate the posterior distribution.

Since the true COVID-19 incidence in the OSU on-campus cohort is not known, in order to have a point of comparison to the IDSA model and ascertain its bias correction magnitude (see Section 3.1), we used a smoothed version of the daily positivity rate 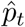. We then computed an associated marginal standard error by 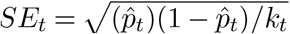 where *k*_*t*_ is the number of tests administered on day *t*. We used these empirical positivity rates 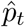 along with their corresponding *SE*_*t*_ to naively estimate at each *t* = 1, …, *T* = 60 the marginal 95% confidence band for the daily prevalence. The results are shown in Figure 6 where in the left panel the median and 2.5% and 97.5% quantiles of the posterior estimates of the OSU daily prevalence are plotted (the blue region) along with the naive marginal estimate of prevalence (based on the values of 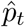) and its 95% confidence band envelope (the pink region).

**Fig 6:**
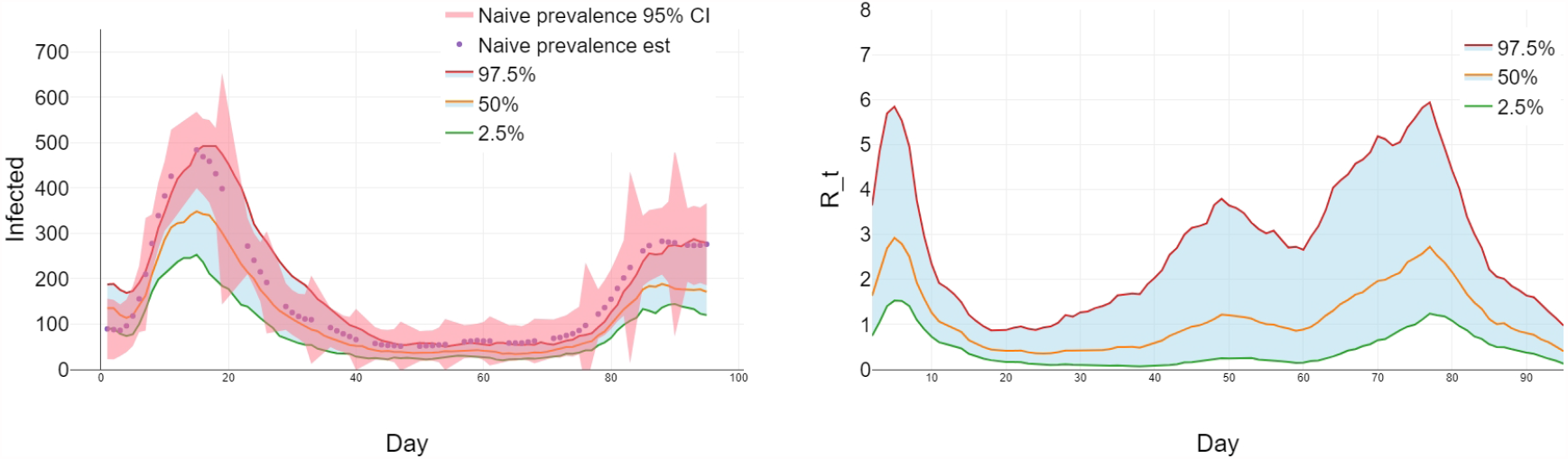
IDSA model fit to OSU COVID-19 data. *Left:* Model estimated prevalence (smooth lines within the blue 95% credibility envelope) vs the naive estimate of prevalence (purple dots within the pink 95% confidence envelope). The IDSA estimates are seen to adjust downward the naive estimates, in accordance with the discussion in Section 3.1. *Right:* The model-based posterior estimate of the effective reproduction number ℛ_*t*_ given by (5). The median and 2.5% and 97.5% quantiles of the posterior estimates are plotted as smooth solid curves within the blue 95% region of credibility.

While the 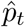’s are somewhat useful as points of comparison, the discussion in Section 3.1 suggests that they have a positive bias that is largest at the epidemic peaks. Indeed, from the plot in the left panel of Figure 6 we see the IDSA fit matches the shape of the epidemic suggested by the 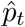, but estimates the peak of the epidemic are at a much lower value. This is consistent with the synthetic data results from previous sections where IDSA is seen to correct the positivity bias described in Section 3.1. The exact amount of this bias present in the OSU data is an area for future work, as the OSU testing scheduling scheme is more complicated than the one discussed in Section 3.1(see Figure 1).

## 4. Conclusions

In the absence of an easily available vaccine, the ability to detect and isolate infections as they occur is essentially the only way to prevent large outbreaks in residential populations such as that of undergraduate students living on a college campus. Assuming that infections among residents may be largely non-symptomatic, the repeated testing scheme needs to be employed to quickly remove active infections and to identify the underlying parameters of disease spread, such as the effective reproduction number, in order to plan for the isolation and quarantine needs. We have presented here a statistical IDSA model that provides such estimates for prevalence and removal rates as well as for the effective reproduction number. The model fits given in Section 3 are encouraging both in synthetic data examples as well as when analyzing OSU repeated testing data. We note that ability of the IDSA model to flexibly fit the changing epidemic pattern largely stems from the fact that we allow for the key transmission (*β*_*t*_) and recovery (*γ*_*t*_) parameters to vary with time. Despite that, the model is fully identifiable as long as the volume of daily and weekly data available for model fitting is sufficiently high. That is clearly the case for the OSU dataset where each week of repeated testing adds on the order of 12000 new data points for parameter estimation. Although not discussed here, in our experience with the IDSA model applied to OSU dataset we have seen good agreement between model forward predictions and the actual data, alleviating the overfitting concerns associated with similar complex mechanistic models.

The analysis of the OSU dataset presented in this paper is to our knowledge the first detailed statistical study of repeated SARS-CoV-2 testing data from a large US college undergraduate setting. For the OSU analysis, we have used the IDSA closed model described in Section 2.2, which in particular neglects importation of infection from outside the campus. Because of this, caution should be exercised in interpreting our ℛ_*t*_ estimates for OSU in the temporal regions of low prevalence. For example, steady low-level importation of cases could make ℛ_*t*_ appear to be around one, despite little transmission within the residential undergraduate community. In our applied work for the University, to account for this we have expanded the model to include a constant baseline level of positive tests per day. This baseline level could correspond to false positives, external imported infections, or a combination of the two. Separating these baseline positives into false positives versus imported infections may require additional data, and is an area for future work. Quantifying the importance of transmission between residential and off-campus members of the university community remains an open question, and one that we hope to use the open model in Section 2.6 to address in the near future with access to a more complete dataset.

We note that as more residential surveillance data is becoming available, further extensions of the open model are also possible and the brief outline of the approach to this end is given in the appendix. However, even though such more general models have the ability to adjust for different types of interactions and social mixing *between* groups, they still may not properly account for the social networks of contacts *within* different groups. Thus additional work is needed for examining SARS-CoV-2 dynamics in the relevant on- and off-campus populations, in order to further improve the accuracy of the IDSA model-based predictions. This is left for future investigation as part of the expansion of the framework presented here. Despite these limitations, we hope that the presented framework will be useful in general and also in the specific context of introducing and evaluating different intervention types including also various vaccination schemes for residents along the lines of some recent policy suggestions Wang et al. (2021). The vaccination issues specifically appear to be quite relevant and could require some adjustments of our modeling methodology. This will be investigated in detail in our future work on IDSA.

## Data Availability

All datasets from Sec 3.3-4  considered in the paper, along with an additional synthetic dataset inspired by the OSU student testing data and the python implementation of the parameter estimation algorithms, are provided at the link below.

https://github.com/mwwascher/IDSA

## Acknowledgements

MW was supported by NSF RAPID Grant DMS-2027001. WRK was supported by the President’s Postdoctoral Scholars Program (PPSP) of the Ohio State university. GAR was supported by NSF RAPID Grant DMS-2027001 and NSF Grant DMS-1853587. PMS was supported by the NIH Grant UL1TR002733. The authors are grateful to Yuhan Pan, Namrata Banerji, Steve Chang, Ron Davies, and Tanya Berger-Wolf for support with data acquisition and cleaning. This project was partially supported by the Ohio State University’s Comprehensive Monitoring Team.

## Ethics Statement

The student testing data for adults only (students 18 years old or older at the time of first testing) were de-identified and extracted from The Ohio State University operational screening for SARS-CoV-2 under research protocol 2021H0189, approved by The Ohio State University Institutional Review Board.

## SUPPLEMENTARY MATERIAL

### GithHub Repository for Data and Analysis Scripts

The datasets in Sections 3.3-3.4, along with an additional synthetic dataset inspired by the OSU student testing data and the python implementation of the parameter estimation algorithms, are provided at https://github.com/mwwascher/IDSA.

## APPENDIX A

### EXTENSION TO *k*-GROUPS MODEL

We may also extend the model in Section 3.4 to an arbitrary number *k* of groups. As our main consideration remains a college campus model, we assume that the first *k* − 1 groups are different on-campus populations under repeated testing while the *k*th group being the background (off-campus) group as in Section 3.4. This model might apply in a situation where several on-campus groups, for example students, faculty, and staff, are all under repeated (possibly different) testing regimes and with different socially behaviors.

Our proposed model for this situation is as follows. Each of the *k* groups is divided into SEIR compartments as before. Denoting the total number of individuals in each group *n*_1_ … *n*_*k*_ respectively, the number of susceptible individuals in each group at time *t* by 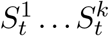 respectively, the number of infected individuals in each group at time *t* by 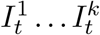 respectively, and so on. We assume *k ≪ n*_*i*_ for *i* = 1 … *k*. The compartment counts then evolve according to the following rules.

- Each pair of one individual among 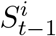 and one among 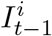 has probability 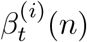 of contact, and each individual in 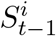 who experiences such a contact is infected/exposed beginning at *t*
- Each pair of one individual among 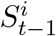 and one among 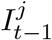 such that *i ≠ j* has probability 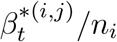 of contact, and each individual in 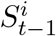 who experiences such a contact is infected/exposed beginning at *t*.
- Following exposure, an individual enters the corresponding exposed compartment. Each individual in 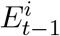 moves to the corresponding *I* compartment at time *t* with probability *ψ* and becomes infectious.
- Each infectious individual in 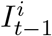 is removed beginning at *t* with probability 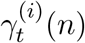.

Let 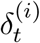 be the daily decrease in susceptibles for group *i*, and let 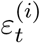 be the daily decrease in infectious for groups *i* for each *i* = 1 … *k*. The probability laws for these quantities are given by^3^

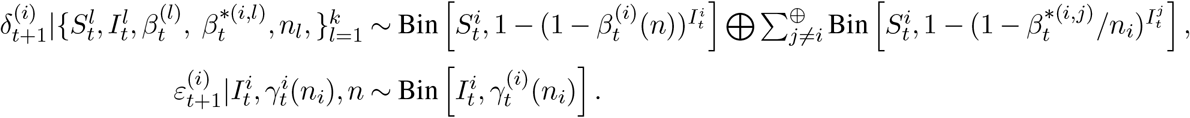

Define 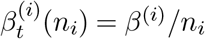,and 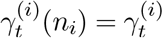 for each *i* = 1 … *k*. When the *n*_*i*_’s are large we can approximate the laws of the 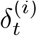 using Le Cam’s theorem by

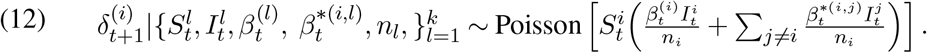

If we now let 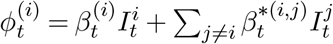 then we can rewrite (12) as

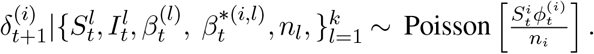

With the help of these formulae one can now build in a relatively straightforward manner an extension of the MH-within-Gibbs sampler algorithm discussed in Section 2.8.

#### A.1. Survival functions for *k*-groups

Let 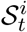 be the survival functions that describe the decay of susceptibles for group *i* for each *i* = 1 … *k*. Conditional on 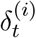, each 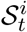 can be described by the recursive equation

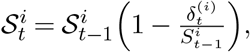

and so the marginal distribution is given by

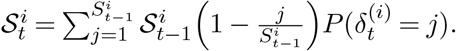

which is a straightforward extension of the formula (9). Incorporating the *E* compartment is now done as in (10).

## APPENDIX B

### FULL CONDITIONAL DISTRIBUTIONS FOR THE GIBBS SAMPLER

#### B.1. Full conditional distribution for *δ*

We obtain values of 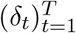 based on a random sample of individual infection times. For individual *i*, let *W*_*i*_ be the time of infection (exposure) and let *X*_*i*_ ∼Geom [*ψ*] be the time spent in the exposed (infected but not yet infectious) state. Then for individual *i*, conditionally on *Z*_*i*_ = *W*_*i*_ + *X*_*i*_ and other random quantities present in the model, we have

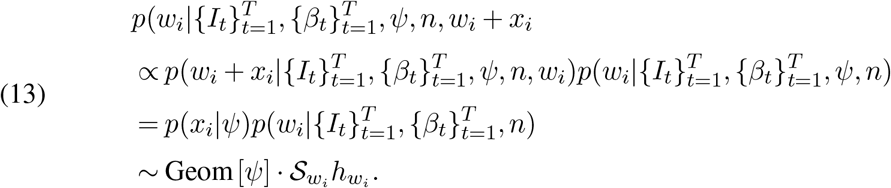

To draw *w*_*i*_ from its full conditional, we use the Metropolis-Hastings step. Given a prior draw *w*_*i*_, we make the next draw as follows.

1. Propose 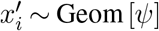.
2. Compute 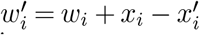.
3. Accept 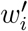 with probability

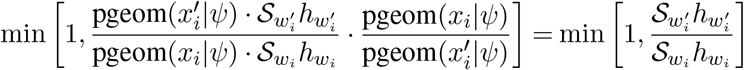

where pgeom(*x*|*ψ*) is a shorthand for *P* (*X* = *x*) when *X* ∼ Geom[*ψ*]. Since the vector 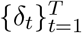 gives the daily counts of newly exposed individuals, it can be now obtained by simply daily aggregation of the infection (exposure) times *w*_*i*_.

#### B.2. Full conditional distribution for *ε*

Let *ε*_*i*_ be the number of individuals removed from the infectious (*I*) compartment between time *i –* 1 and *i*. Then 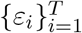 denotes all the removal counts. Then, conditionally on the remaining random quantities

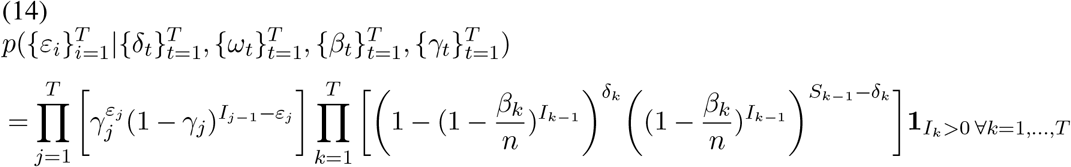

Note that 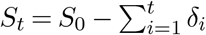 and 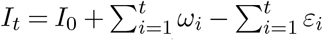 can be computed from the other quantities and are used in equation (14) to make it more readable.

To make draws of 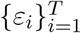 from its full conditional we use as above the Metropolis-Hastings step. Given a prior draw of 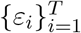, we make the next draw as follows.

1. Propose 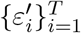 by proposing each 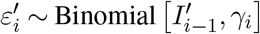
2. Compute 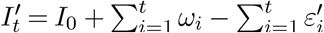
3. Accept 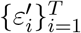 with probability

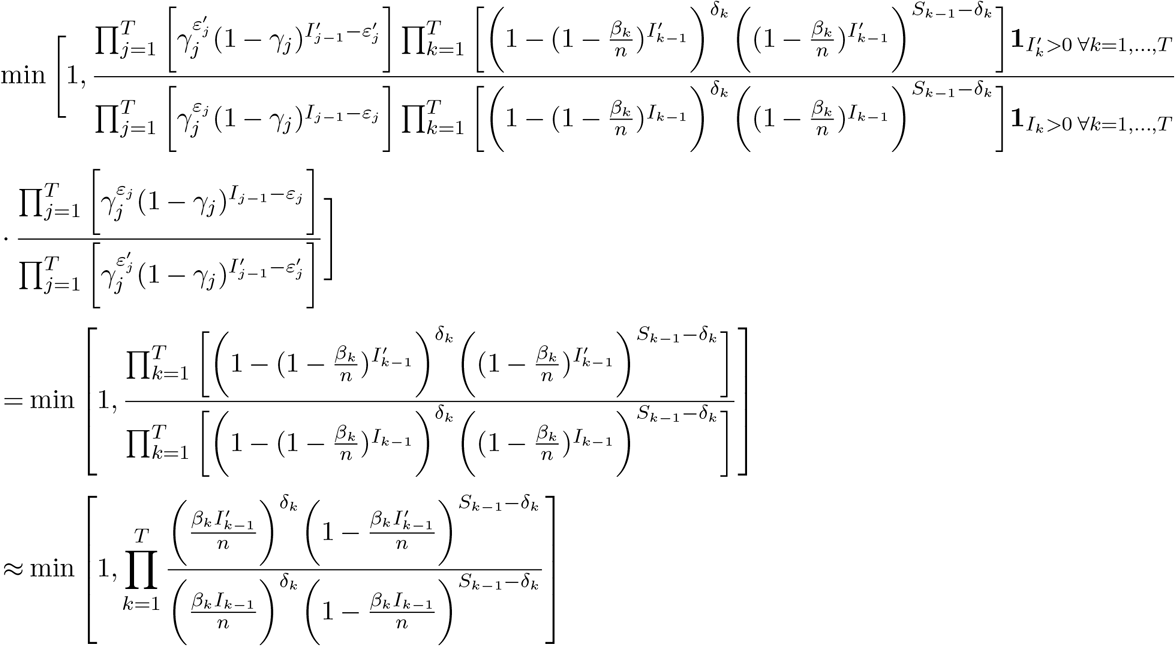

It is possible but in practice highly unlikely that this ratio could be 0*/*0 and thus undefined. If this happens, we accept 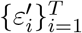. A sequence 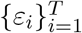 can have likelihood of 0 if it would cause *I*_*t*_ = 0 while *δ*_*t*_ > 0, since this would imply infections occurred while no individuals were infectious. A proposed 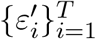 could cause this if on some day *t* where *δ*_*t*_ > 0 we observe that 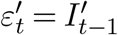 and *ω*_*t*_ = 0 so that all infectious individuals at time *t* − 1 are removed and no exposed individuals at time *t* − 1 more to the infectious compartment. In practice this is highly unlikely. A retained sequence 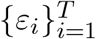 can cause this because we update 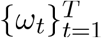 using an approximation and so do not check the compatibility of this sequence with 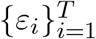 in the updating step. Thus, it is possible that updating 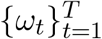 and retaining 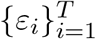 could cause *I*_*t*_ = 0 while *δ*_*t*_ > 0. Again, in practice we found this 0*/*0 case is highly unlikely and almost never occurs.

## APPENDIX C

### SUMMARY OF NOTATION

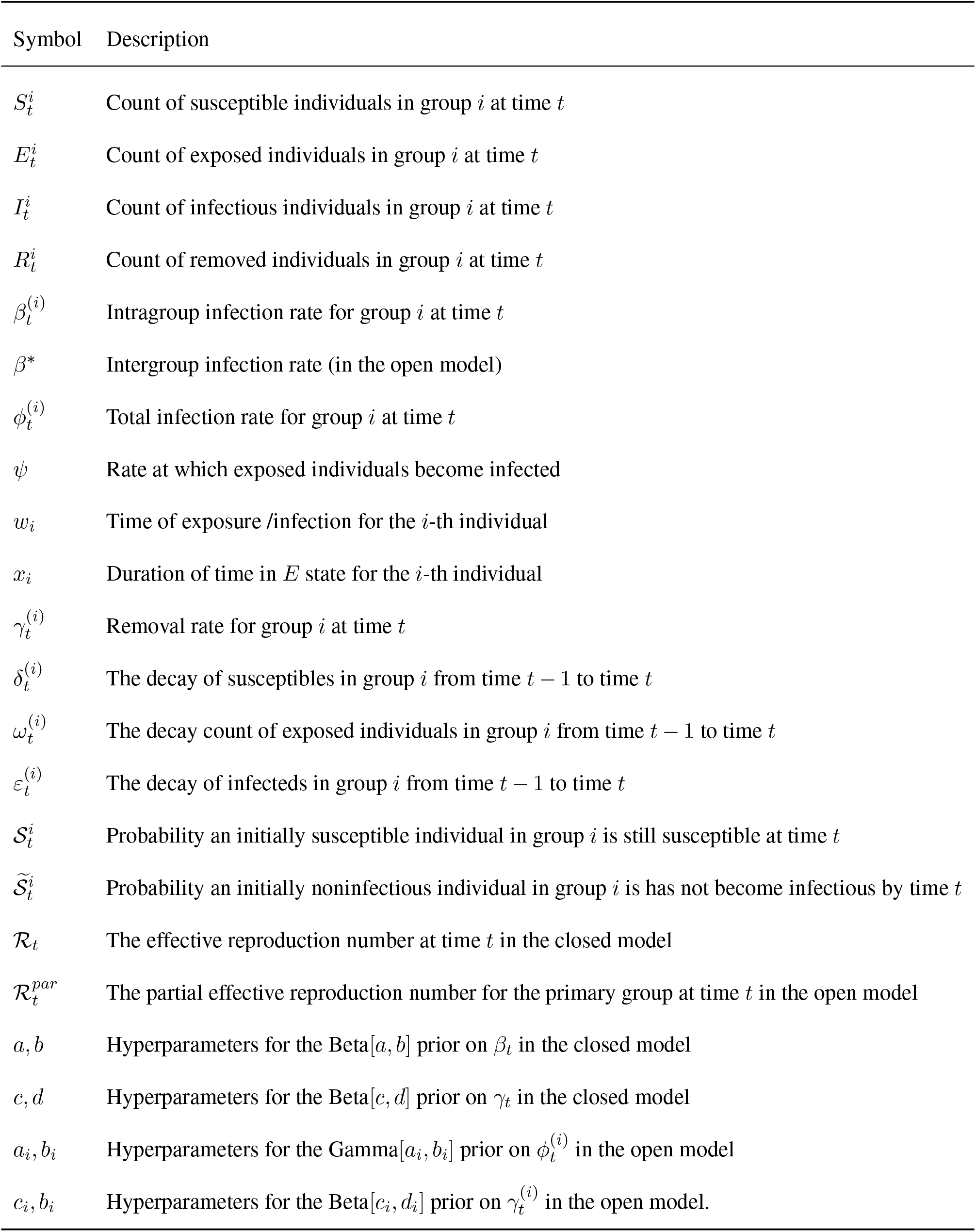

APPENDIX C: SUMMARY OF NOTATION
| Symbol | Description |
| --- | --- |
| $S_t^i$ | Count of susceptible individuals in group $i$ at time $t$ |
| $E_t^i$ | Count of exposed individuals in group $i$ at time $t$ |
| $I_t^i$ | Count of infectious individuals in group $i$ at time $t$ |
| $R_t^i$ | Count of removed individuals in group $i$ at time $t$ |
| $\beta_t^{(i)}$ | Intragroup infection rate for group $i$ at time $t$ |
| $\beta^*$ | Intergroup infection rate (in the open model) |
| $\phi_t^{(i)}$ | Total infection rate for group $i$ at time $t$ |
| $\psi$ | Rate at which exposed individuals become infected |
| $w_i$ | Time of exposure /infection for the $i$-th individual |
| $x_i$ | Duration of time in $E$ state for the $i$-th individual |
| $\gamma_t^{(i)}$ | Removal rate for group $i$ at time $t$ |
| $\delta_t^{(i)}$ | The decay of susceptibles in group $i$ from time $t - 1$ to time $t$ |
| $\omega_t^{(i)}$ | The decay count of exposed individuals in group $i$ from time $t - 1$ to time $t$ |
| $\varepsilon_t^{(i)}$ | The decay of infecteds in group $i$ from time $t - 1$ to time $t$ |
| $\mathcal{S}_t^i$ | Probability an initially susceptible individual in group $i$ is still susceptible at time $t$ |
| $\tilde{\mathcal{S}}_t^i$ | Probability an initially noninfectious individual in group $i$ is has not become infectious by time $t$ |
| $\mathcal{R}_t$ | The effective reproduction number at time $t$ in the closed model |
| $\mathcal{R}_t^{par}$ | The partial effective reproduction number for the primary group at time $t$ in the open model |
| $a, b$ | Hyperparameters for the Beta$[a, b]$ prior on $\beta_t$ in the closed model |
| $c, d$ | Hyperparameters for the Beta$[c, d]$ prior on $\gamma_t$ in the closed model |
| $a_i, b_i$ | Hyperparameters for the Gamma$[a_i, b_i]$ prior on $\phi_t^{(i)}$ in the open model |
| $c_i, d_i$ | Hyperparameters for the Beta$[c_i, d_i]$ prior on $\gamma_t^{(i)}$ in the open model. |

## Footnotes

1 Here ⊕ indicates that we take a sum of the independent random variables.

2 Students who were minors were removed from our analysis as per institutional policy.

3 Here the notation ⊕ and ∑^*⊕*^ indicates that we are considering distributions obtained by summing independent random variables.

